# Physicians’ Industrial Action: An Interrupted Time Series Analysis Assessing Impact in 11 Kenyan Neonatal Units

**DOI:** 10.64898/2026.10.07.26364929

**Authors:** Meghan Paral, Christine Manyasi, John Wainaina, George Okello, Dolphine Rabera Mochache, Hannah Mwaniki, Domitilla Mueni Muinde, Franklin Ochieng Okech, Joy E Lawn, Natasha R Rhoda, Rebecca Richards-Kortum, Z Maria Oden, Lisa R Hirschhorn, Christine A Bohne, William M Macharia

**Affiliations:** Rice360 Institute for Global Health Technologies, Rice University, Houston, Texas, USA; Department of Paediatrics, Aga Khan University, Nairobi, Kenya; Feinberg School of Medicine, Northwestern University, Chicago, USA; Children’s Institute, Dept. Paediatrics and Child Health, Faculty of Health Sciences, University of Cape Town, Cape Town, South Africa; KEMRI-Wellcome Trust, Nairobi, Kenya; Mbagathi County Hospital, Nairobi, Kenya; London School of Hygiene and Tropical Medicine, London, UK

**Keywords:** Small and sick newborn care, Neonatal care, Industrial action, Healthcare worker strikes, Health systems shocks

## Abstract

**Background:** Skilled healthcare workers are essential for neonatal survival; however, the Kenyan public health system has historically faced strikes compounded by a human resource crisis. Evidence on how strikes affect neonatal care and mortality remains limited. This study evaluated changes in neonatal admissions, clinical care processes, and inpatient mortality associated with 56 days of disruption during the 2024 Kenyan physicians’ strike.

**Methods:** We conducted a retrospective interrupted time-series analysis of 12,184 admissions to 11 Kenyan neonatal units implementing with the NEST360 Alliance, recorded in the Neonatal Inpatient Dataset from December 2023 to August 2024. Weekly admissions and inpatient mortality were modeled over a 72-week window (27,638 admissions) using hospital-clustered generalized estimating equations with seasonal terms. Care processes, case fatality, and birthweight-specific mortality were compared across study periods.

**Results:** Weekly admissions fell from 413 to 189 during the strike (incidence rate ratio (IRR) 0.49, 95% confidence interval (CI) 0.31–0.75), approximately 1,700 fewer admissions than expected, and returned to baseline afterward (IRR 1.00, CI 0.81–1.23). Newborns admitted during the strike were of lower birthweight, more often premature, and more often outborn. Admissions fell across all birth-location categories, most steeply among inborn neonates. No measurable change was detected in documented inpatient coverage of care measures, in crude mortality (12.5% to 14.9%; risk ratio (RR) 1.07, CI 0.85–1.33), or in trend-adjusted mortality (IRR 0.88, CI 0.53–1.45). Case fatality among neonates with recorded infection doubled, from 4.7% to 9.6% (RR 2.06, CI 1.18–3.58; adjusted p = 0.047).

**Conclusions:** The 2024 physicians’ strike was associated with severe disruption to neonatal service utilization, with admissions returning to baseline when the strike ended. Among newborns who continued to reach care, inpatient mortality did not change measurably, but case fatality among those with recorded infection doubled. This analysis was confined to inpatient data; hence, the outcomes of newborns who did not reach the hospitals were not captured. Both inpatient and population-level data are needed to assess the full impact of the strike. Mitigating harm during future labor disruptions requires interventions such as resilient care models, formalized task-shifting protocols, and minimum service coverage agreements.

**WHAT WAS KNOWN?:**

- The public health care system in Kenya experiences recurrent healthcare worker strikes during which maternal and general health services utilization is disrupted
- Neonates are among the most vulnerable populations in the Kenyan public health system. However, few publications have quantified the impact of healthcare provider strikes on neonatal outcomes and processes

**WHAT WAS DONE THAT IS NEW?:**

- Data from 11 Kenyan neonatal units, comprising 12,184 admission records from December 2023 to August 2024, were analyzed to evaluate changes in neonatal admissions, case mix, care processes, and inpatient mortality associated with a 56-day national physicians’ strike
- Because the strike had an exact-dated onset and conclusion, both the disruption and recovery were estimated using an interrupted time series design, with hospital-clustered generalized estimating equations applied across all 11 units

**WHAT WAS FOUND?:**

- Weekly admissions fell by approximately half during the strike, an estimated shortfall of around 1,700 admissions against seasonal expectations, and returned to seasonal baseline once the strike concluded
- Newborns admitted during the strike period were of lower birthweight, more often premature, and more often outborn. Admissions fell across every birth-location category but most steeply among neonates born in the admitting hospital
- There was no measurable change detected in documented coverage of seven screening and intervention measures, or in inpatient mortality overall or by birthweight band. Case fatality among neonates with infection as the primary discharge diagnosis doubled.

**WHAT NEXT?:**

- Inpatient data do not capture the outcomes of newborns who failed to access hospitals. Community-level data are necessary to quantify the full impact of service disruption.
- As industrial actions continue to be increasingly prevalent with frequent disruption of services, mitigation strategies should be put in place to protect vulnerable patients.

## Background

Kenya’s public hospitals experience recurrent workforce strikes, driven by chronic health worker shortages, uneven post-devolution labor conditions, and unresolved compensation disputes [1–5]. These disruptions afflict a system already struggling to protect its most vulnerable patients: newborns account for 46% of pediatric admissions nationally, but 66% of inpatient pediatric deaths [6], and Kenya remains off track to meet the Sustainable Development Goal (SDG) target of 12 or fewer neonatal deaths per 1,000 live births by 2030 [6].

Neonatal units are poorly positioned to absorb physician absence. Even under routine conditions, Kenyan public neonatal units operate at median nurse-to-newborn ratios ranging from 1:13 to 1:25, far exceeding the ratios recommended in national guidelines, limiting the time available per newborn and contributing to missed care [7–10]. Within the standard clinical hierarchy, physicians lead diagnostic and treatment planning, while nurses implement care. Experienced nurses can independently sustain routine interventions for stable newborns, but complex or deteriorating cases require physician-led decision-making [11]. A physicians’ strike would therefore be expected to disproportionately compromise care for the sickest and most complex neonatal admissions.

Prior studies of Kenyan strikes document broader consequences such as missed antenatal-to-postnatal care, reduced detection of complications, and minimal substitution by private or faith-based providers during the 2016–2017 strikes [12, 13]. However, literature has focused on maternal and general health service utilization. Data quantifying strike-related changes in neonatal-specific care processes and outcomes are limited.

Between March 15 and May 9, 2024, Kenya experienced a 56-day nationwide physicians’ strike involving roughly 4,000 doctors, nearly half the country’s registered physician workforce [14, 15]. The Newborn Essential Solutions and Technologies (NEST360) alliance provides an opportunity to evaluate neonatal admissions and outcomes during this strike. NEST360 is an international alliance of 23 institutions and organizations working with governments in five African countries (Ethiopia, Kenya, Malawi, Nigeria, and Tanzania) to improve hospital newborn care through appropriate technologies, workforce skill-building, and data for quality-improvement strategies. As of September 2025, two-thirds of the 65 hospitals supported by NEST360 had reductions in mortality, and 40% of hospitals had significant reductions ranging from 13–50% [16]. Routine admission data collected in partnership with the NEST360 alliance pose a unique opportunity to study specific impacts of system shocks on neonatal care. Similar work has previously been done in the assessment of disruptions due to the COVID-19 pandemic [17].

Using an interrupted time series (ITS) design, this study assessed whether and how the 2024 national physicians’ strike affected neonatal units implementing NEST360 in 11 Kenyan public hospitals. Specific objectives were to evaluate changes in (1) neonatal admissions, (2) care practices including kangaroo mother care (KMC), phototherapy, continuous positive airway pressure (CPAP) use, and screening for glucose, oxygen saturation, and temperature at admission, and (3) inpatient outcomes, including overall mortality and diagnosis- and birthweight-specific case fatality.

## Methods

### Study Design and Setting

We conducted a retrospective, multicenter ITS analysis [18] of 11 of the 13 Kenyan public hospitals implementing with the NEST360 Alliance, each with a single neonatal unit [19]. Two were excluded because of unrelated disruptions during the pre-strike period that could confound results. One unit is a tertiary teaching referral hospital providing WHO Level III neonatal care; the remaining ten provide Level II care, including CPAP.

The interruption event was a 56-day nationwide physicians’ strike spanning March 15 to May 9, 2024. The study timeline was divided into three periods for descriptive analysis: a three-month pre-strike baseline (December 15, 2023–March 14, 2024), the strike period (March 15–May 9, 2024), and a three-month post-strike period (May 10–August 9, 2024). Because the strike had a defined end date, both onset and recovery effects were modeled.

Since annual seasonality cannot be identified within a nine-month span, ITS models were estimated over an extended 16-month (72-week) window (March 20, 2023–August 4, 2024), ending with the last complete calendar week. Per-hospital inspection of weekly admissions across this window identified no other hospital-level disruptions (no gaps ≥3 weeks, no admission shifts >2-fold).

### Data Source and Study Population

Data were obtained from the NEST360 Neonatal Inpatient Dataset (NID), a standardized clinical dataset co-developed with partner governments and abstracted from routine neonatal inpatient records by trained data officers through Kenya’s Clinical Information Network (CIN) [20, 21].

The ITS model was informed by weekly admission counts from among 27,638 admissions across the 11 units during the extended analysis window. Forty-four percent (n = 12,184) of these admissions were in the three predefined periods (n = 5,364 pre-strike, 1,512 strike, 5,308 post-strike), forming the cohort for descriptive and subgroup analyses. Admissions (n = 15,454, 55.9%) outside these periods contributed only to the estimation of secular trend and seasonality.

Inpatient mortality was based on recorded discharge status; admissions with unknown or transfer outcomes were retained in admission counts but excluded from mortality denominators (12,175 analyzable admissions; 1,501 deaths). Admission type (inborn/outborn) was derived from birth location, and diagnosis from the primary discharge diagnosis. Outborn admissions were further classified as either referred from another health facility or born before arrival (at home, in transit, or at another non-facility location). Infection as diagnosis was defined as the primary discharge diagnosis as recorded by clinicians. This is a clinical diagnosis with no standard case definition, may not be consistently recorded, and may not be culture-confirmed, given limited blood culture capacity [22].

### Variables and Outcome Definitions

Primary outcomes were weekly neonatal unit admissions and weekly inpatient mortality (deaths per admission with known outcome). Secondary variables included patient characteristics (birthweight, admission type, age at admission), birthweight-specific mortality, diagnosis-specific case fatality, and screening/intervention coverage among eligible neonates, as follows: KMC (1,000–1,999 g); CPAP (1,000–1,499 g, or 1,500–1,999 g with severe respiratory distress, indrawing and grunting); phototherapy (for admissions with documented clinical jaundice); vitals monitored among all admissions at admission (glucose, pulse oximetry, and temperature screening); and admission normothermia (36.5–37.5°C) [23].

To describe the different effects of the strike, units were stratified by the magnitude of admission decline: one unit was denoted minimally reduced as it maintained services via contracted staff, and the remaining ten were split at the median into severely reduced (five units, 55.7–96.5% decline) and moderately reduced (five units, 20.8–50.5% decline) groups. As these groups were defined by the outcome itself, group-level admission, coverage, and case-fatality results are reported descriptively rather than by testing of hypotheses.

### Statistical Analysis

#### Model specification

Data were aggregated to the unit-week level (11 units × 72 weeks = 792 unit-weeks). Weekly admission counts were modeled using Poisson regression with a log link, yielding incidence rate ratios (IRRs) for multiplicative change relative to baseline trend. Segmented regression distinguished abrupt level changes at strike onset from gradual slope changes, with additional terms capturing post-strike recovery (full model output in Additional file 1).

The model took the form:

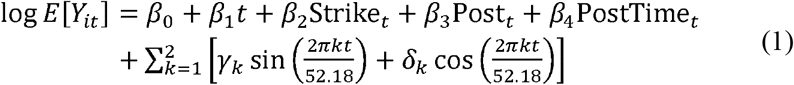

- *Y_it_* is the admission count in neonatal unit *i* during calendar week *t*.
- *β_1_* denotes the baseline weekly trend.
- *Strike_t_* is an indicator variable for strike-period weeks, with *exp*(*β_2_*) representing the primary IRR (strike-period level change).
- *Post_t_* is an indicator variable for post-strike weeks, with *exp*(*β_3_*) representing the post-strike level change.
- *PostTime_t_* denotes the number of weeks elapsed since the strike ended, with *exp*(*β_4_*) reflecting the post-strike slope change per week.
- *γ_k_* and *δ_k_* are harmonic (Fourier) coefficients adjusting for annual seasonality (period = 52.18 weeks, *K=2*, where 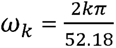) [24].

Mortality was modeled identically, with weekly deaths as the outcome and log(weekly admissions with known outcome) as an offset, scaling estimates to deaths per admission rather than admission volume. Unit-weeks with zero analyzable admissions (n = 6) were excluded, yielding 786 unit-weeks for the mortality model.

Intervention coverage, diagnosis-specific case fatality and birthweight-specific mortality were compared between the pre-strike and strike periods across all 11 units using population-averaged generalized estimating equations (GEE) clustered by hospital [25], with a logistic link for coverage, yielding odds ratios (ORs), and a log link for case fatality and mortality, and the same Mancl–DeRouen correction and t(10) inference as the admission models.

#### Variance and clustering

Models were estimated via population-averaged GEE clustered by neonatal unit, yielding marginal effects across hospitals. Working correlation structure was selected by quasi-likelihood information criterion (QIC) for the admissions and mortality models [26]. An exchangeable structure was pre-specified for coverage and case-fatality models. Robust (sandwich) standard errors were used to address overdispersion, with the Mancl–DeRouen bias-reduced correction applied [27], given the limited number of clusters (11 hospitals) [28]. Confidence intervals (CIs) and p-values used a t-distribution with 10 degrees of freedom.

#### Sensitivity analyses

Robustness was assessed via: (i) a negative binomial GEE modeling extra-Poisson variation directly (maximum-likelihood dispersion α = 0.040); (ii) restriction to the three-month pre-strike baseline, omitting harmonic terms; (iii) temporal anchoring to strike start date rather than calendar week; (iv) a pooled negative binomial specification (distinct from (i)) with Newey–West standard errors; (v) hospital-level segmented models to describe cross-unit heterogeneity, and (vi) a first-order autoregressive (AR(1)) working correlation structure.

#### Multiple testing

Wilson score intervals were used for descriptive proportions [29]. Benjamini–Hochberg (BH) adjustment was applied within three pre-specified families: seven coverage measures, three diagnosis-specific case-fatality comparisons, and five birthweight bands [30]. CIs are unadjusted. Hospital-level estimates and primary time-series models were not corrected. Significance was set at two-sided adjusted p < 0.05. All analyses were performed in Python (v3.12) [31].

## Results

### Study Population and Baseline Characteristics

Records of 12,184 neonatal admissions across the 11 neonatal units were included in the three study periods: 5,364 (44.0%) pre-strike, 1,512 (12.4%) during the strike, and 5,308 (43.6%) post-strike (Table 1). Of 12,175 (99.9%) admissions with a known outcome, 1,501 (12.3%) died. The descriptive window contributed 374 unit-weeks of observation (34 calendar weeks × 11 units). The ITS models were estimated over an extended 72-week window comprising 792 unit-weeks and 27,638 admissions.

**Table 1:**
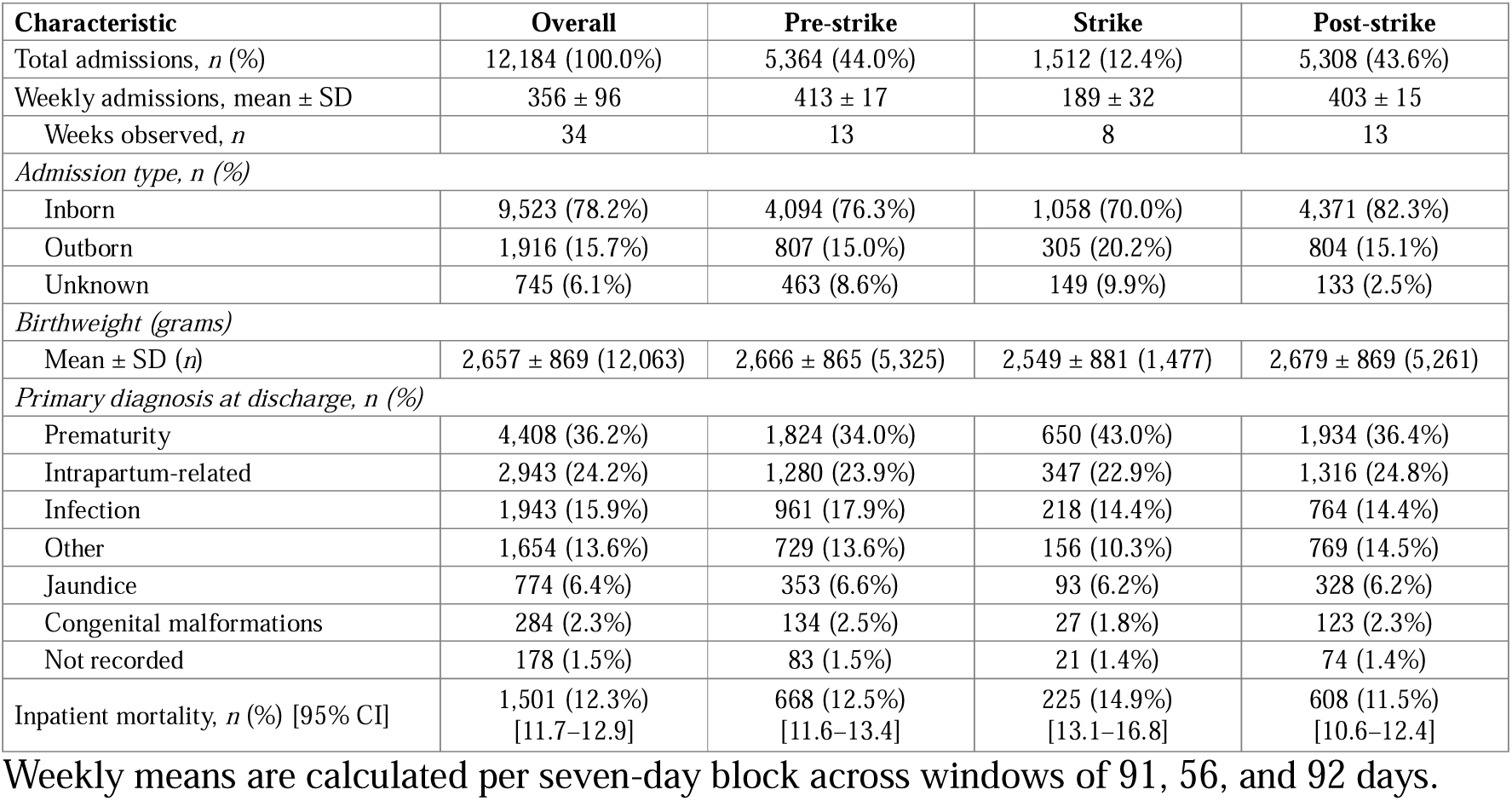
Baseline characteristics across defined strike periods.

Across all 11 neonatal units, admissions during the strike were smaller (mean birthweight 2,666 ± 865 g [mean ± standard deviation (SD)] pre-strike vs 2,549 ± 881 g), more often premature (34.0% vs 43.0%), and more often outborn (16.5% vs 22.4%). Median age at presentation for outborn newborns was both 0 days (interquartile range (IQR) 0–1) pre-strike (n = 787) and during-strike (n = 302). Referrals as a share of outborn admissions decreased from 86.0% pre-strike to 83.3% during the strike and rebounded to 87.3% post-strike. Birth location was unknown for 8.6% of pre-strike and 9.9% of strike-period admissions, but only 2.5% of post-strike admissions.

Patient case-mix shifted most in the severely reduced admissions group, where mean birthweight dropped from 2,677 ± 870 g to 2,138 ± 901 g, admissions with a primary discharge diagnosis of prematurity rose from 31.6% to 54.3%, the outborn share rose from 18.9% to 57.0%, and median length of stay rose from 4 days pre-strike (IQR 2–8 days) to 9.5 days during the strike (IQR 4–25 days). Changes were intermediate in the minimally reduced hospital and minimal in the moderately reduced group (Additional file 2).

### Impact of Strike on Neonatal Admissions

Mean weekly admissions across the 11 units fell from 413 ± 17 per week pre-strike to 189 ± 32 during the strike, a crude decline of 54.2%, and rebounded to 403 ± 15 per week post-strike (Figure 1; Table 1). In the primary ITS model, the strike was associated with an immediate 51.5% reduction in weekly admissions (IRR 0.49; 95% CI 0.31–0.75; p = 0.004). The estimate was significant across all sensitivity specifications, ranging from 0.43 to 0.64: 0.47 (95% CI 0.31–0.74) under the negative binomial GEE, 0.50 (CI 0.31–0.80) restricting to the three-month baseline, 0.43 (CI 0.26–0.71) with strike-anchored weeks, 0.56 (CI 0.47– 0.66) under the original pooled specification, and 0.64 (CI 0.45–0.92) with an AR(1) working correlation.

**Figure 1:**
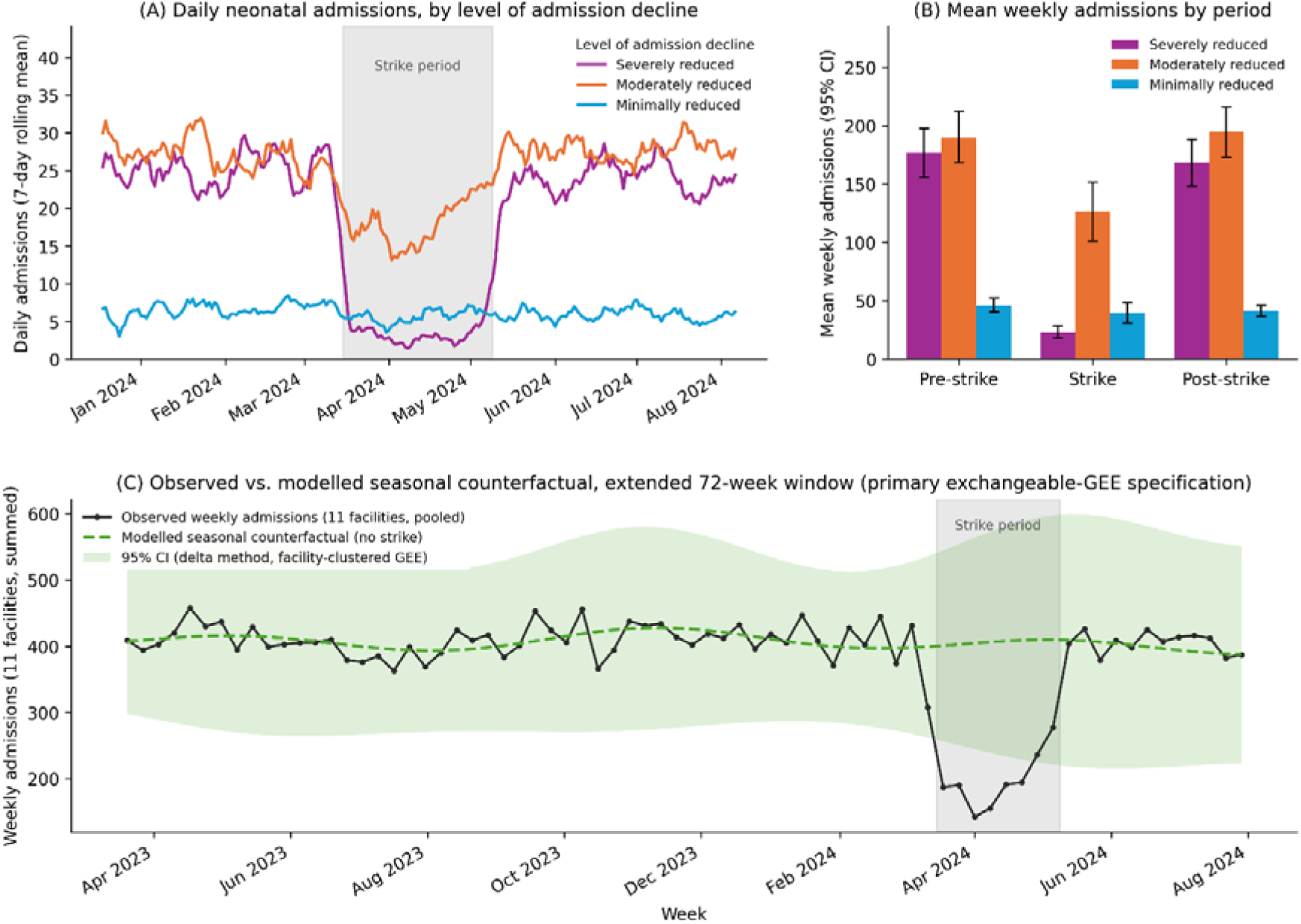
Neonatal admissions from December 2023 to August 2024 in 11 neonatal units in Kenya. (A) Seven-day rolling average of daily admissions stratified by level of reductions in admissions. The gray shaded region indicates the strike period. (B) Mean weekly admissions with 95% CIs across study periods. (C) Observed weekly neonatal admissions (11 hospitals, pooled) against the modeled seasonal counterfactual absent the strike.

As seen in Figure 2, the decline was not uniform across units. Unit-level strike IRRs ranged from 0.05 to 0.93, with a median of 0.57. Eight of the 11 units showed a statistically significant decline. Within the minimally reduced admissions group, mean weekly admissions decreased by 14.6% during the strike, from 46 to 40, and stabilized at 41 post-strike. In the moderately reduced group, admissions fell by 33.5% during the strike, from 190 to 126, and rebounded to 195 post-strike. In the severely reduced group, admissions dropped by 86.8% during the strike, from 177 to 23, and rebounded to 168 post-strike.

**Figure 2:**
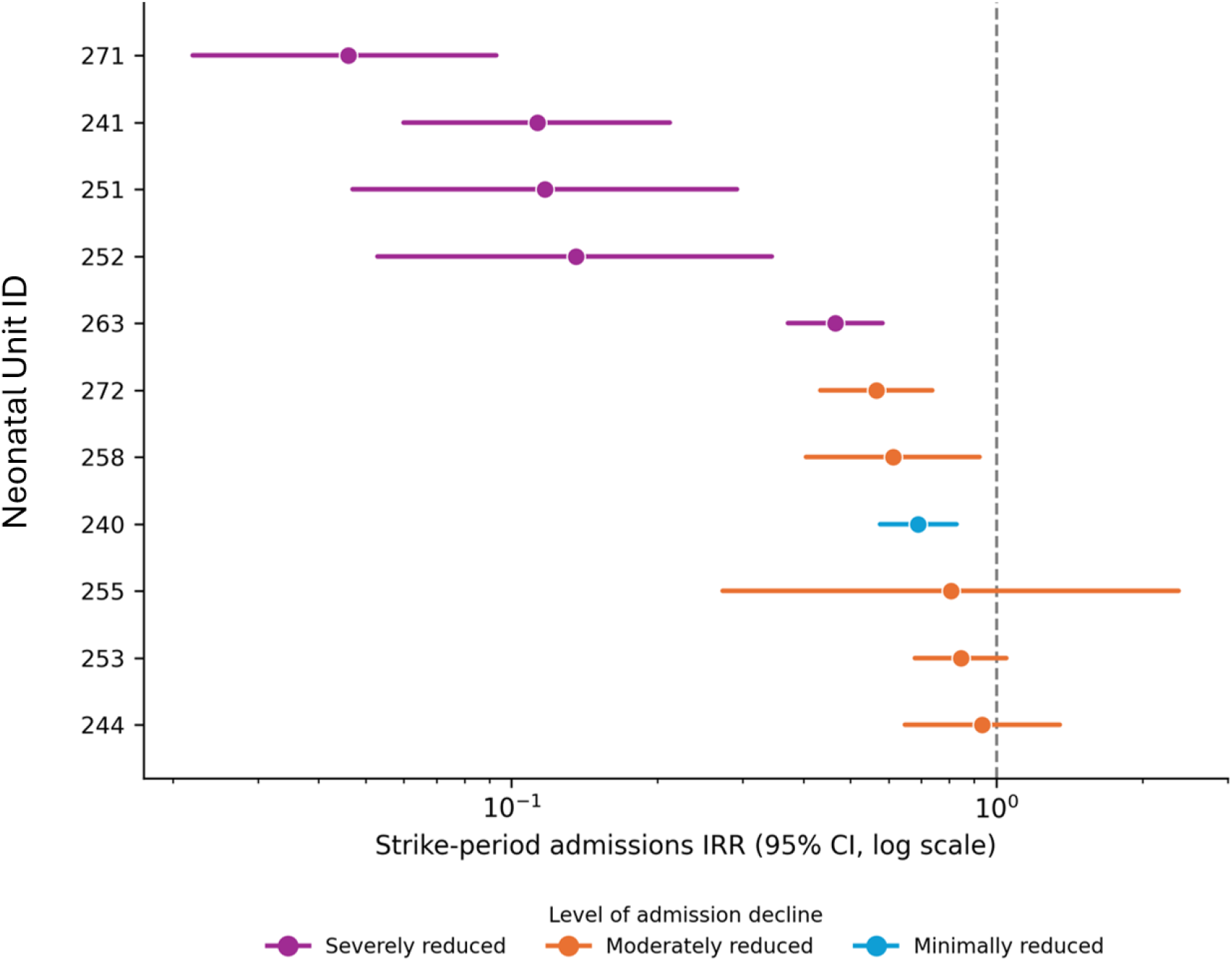
Strike-period IRR for weekly admissions, by neonatal unit. Points are unit-specific estimates from segmented models with 95% CIs; the dashed line marks no change. Units are ordered by point estimate.

Compared with the admissions expected from each unit’s own seasonal baseline, an estimated 3,251 admissions (95% CI 1,901–4,601) would have occurred during the strike weeks in the absence of the strike, against 1,572 observed in the model, a shortfall of 1,679 admissions (95% CI 329–3,029). The post-strike level did not differ from the seasonal baseline (IRR 1.00; 95% CI 0.81–1.23; p = 0.99). There was no residual post-strike trend (IRR 1.00 per week; 95% CI 0.99–1.01). The ITS panel uses complete calendar weeks and therefore includes 1,572 strike-period admissions while the descriptive window (Table 1) uses the exact 56-day strike period, 1,512.

Admissions fell across every birth-location category. Weekly inborn admissions fell from 315 to 132 (−58.0%), referrals from other health facilities fell from 53 to 32 (−40.5%), and births before arrival (outborn non-referrals) from nine to six (−26.7%). Both outborn subcategories rose as a share of admissions with birth location recorded: referrals from 14.2% (95% CI 13.2–15.2) to 18.6% (95% CI 16.7–20.8), and births before arrival from 2.3% (95% CI 1.9– 2.8) to 3.7% (95% CI 2.9–4.9).

### Changes in Care Processes

Across all 11 units, care processes did not change significantly between the pre-strike and strike periods after correction for multiple comparisons (Table 2). Descriptively, the severely reduced admissions group experienced the largest changes in coverage during the strike period, with glucose screening increasing from 84.9% to 93.5%, pulse oximetry increasing from 78.8% to 97.3%, and temperature screening increasing from 93.2% to 96.8% (N = 2,294 pre-strike, N = 186 during the strike). Among eligible newborns, KMC and phototherapy also saw increases from 61.6% (N = 490) to 79.0% (N = 81) and 81.6% (N = 343) to 96.0% (N = 50), respectively, while CPAP coverage declined from 25.2% (N = 250) to 23.8% (N = 42) (Additional file 3).

**Table 2:**
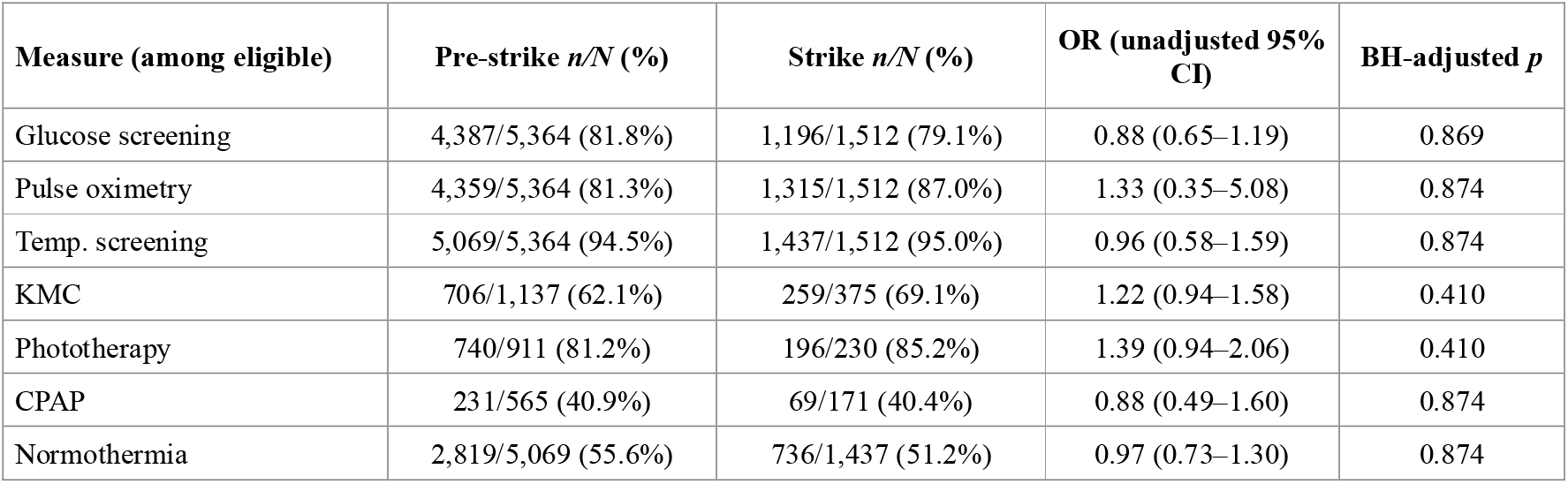
Care process coverage among eligible admissions.

### Impact on Patient Outcomes

Among neonates with infection as the primary diagnosis at discharge, the case-fatality rate across all 11 units rose from 4.7% (45/960) before the strike to 9.6% (21/218) during the strike (risk ratio (RR) 2.06, 95% CI 1.18–3.58; BH-adjusted p = 0.047) (Figure 3). Case fatality was not affected by prematurity and intrapartum-related conditions. Group-level case fatality by diagnosis is reported in Additional file 4.

**Figure 3:**
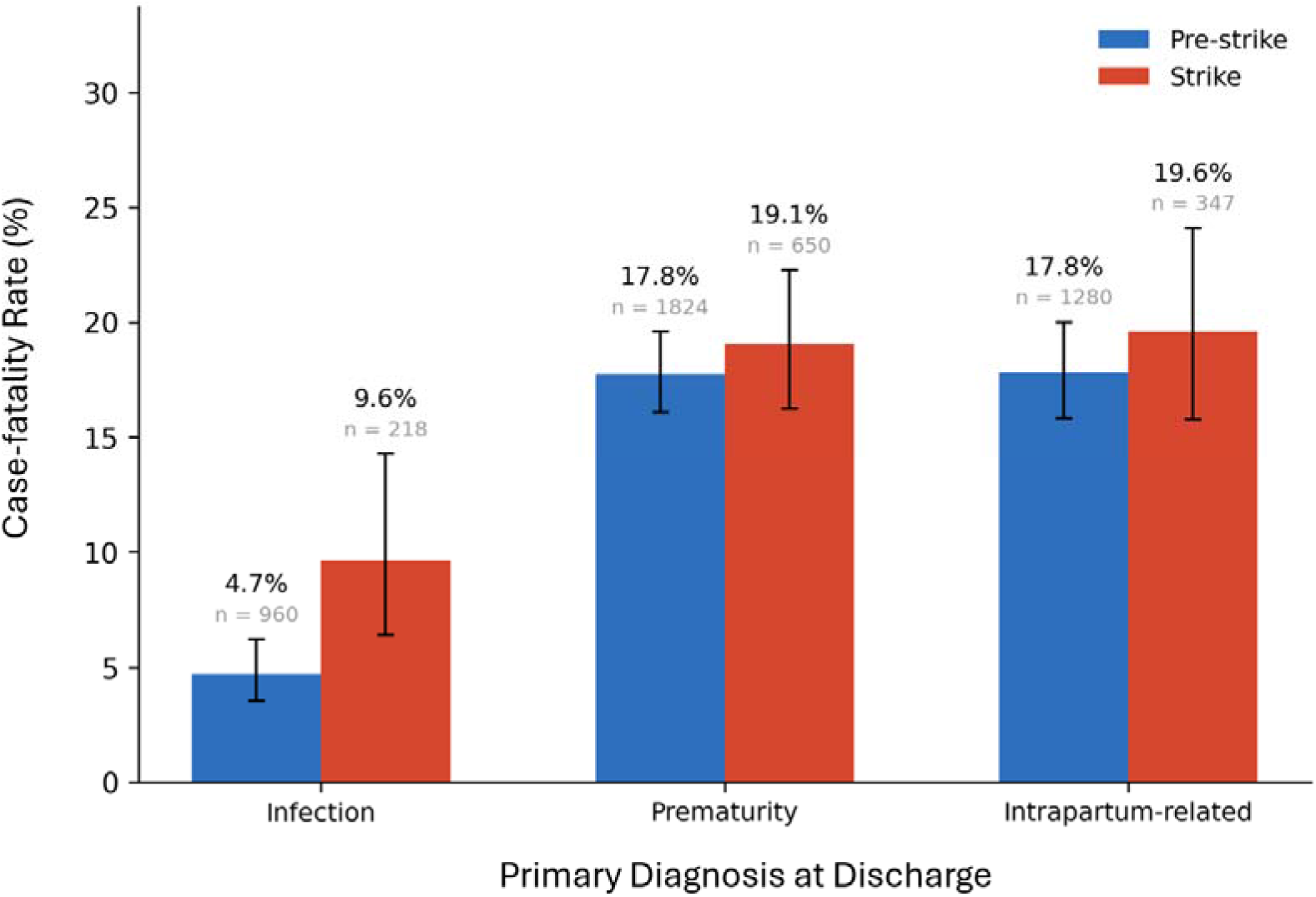
Case fatality by primary diagnosis at discharge, pre-strike and strike periods, across 11 neonatal units.

No birthweight band had significant changes in mortality risk during the strike period (Additional file 5). Mortality among neonates under 1,000 g was 797 and 944 per 1,000 in the two periods, while mortality among neonates over 2,500 g was 75 and 97 per 1,000.

Across all 11 units, crude inpatient mortality was 12.5%, 14.9%, and 11.5% before, during, and after the strike, respectively. Accounting for clustering by hospital, mortality risk did not differ between the strike and pre-strike period (RR 1.07, 95% CI 0.85–1.33; p = 0.54; Table 3). After each unit’s mortality rate was adjusted for its own secular trend and seasonality, the strike-period incidence rate ratio was 0.88 (95% CI 0.53–1.45; p = 0.57).

**Table 3:** Inpatient mortality.

|  | Pre-strike n/N (%) | Strike n/N (%) | RR / IRR (95% CI) | BH p-val |
| --- | --- | --- | --- | --- |
| *Overall mortality* |  |  |  |  |
| Crude | 668/5,360 (12.5%) | 225/1,512 (14.9%) | 1.07 (0.85–1.33) | 0.539 |
| Trend-adjusted | – | – | 0.88 (0.53–1.45) | 0.574 |
| *By birthweight band* |  |  |  |  |
| 400–999 g | 110/138 (79.7%) | 51/54 (94.4%) | 1.18 (1.02–1.37) | 0.163 |
| 1,000–1,499 g | 141/395 (35.7%) | 47/146 (32.2%) | 0.90 (0.63–1.27) | 0.506 |
| 1,500–1,999 g | 95/742 (12.8%) | 28/229 (12.2%) | 0.76 (0.45–1.29) | 0.412 |
| 2,000–2,499 g | 62/758 (8.2%) | 11/192 (5.7%) | 0.69 (0.31–1.55) | 0.412 |
| ≥2,500 g | 246/3,288 (7.5%) | 83/856 (9.7%) | 1.21 (0.97–1.51) | 0.195 |

CI's are unadjusted. Trend-adjusted is the IRR for the strike period of the primary segmented Poisson GEE.

The timing of death remained similar, with 70.2% of inpatient deaths occurring within two days of admission before the strike and 71.1% during the strike.

## Discussion

This study evaluated the impact of a 56-day nationwide physicians’ strike across 11 Kenyan hospitals, observing a 54.2% overall decline in neonatal admissions. In severely reduced admissions hospitals, this reduction was accompanied by a demographic shift toward lower-birthweight and outborn newborns, indicating a concentration of complex and high-risk cases among those who successfully accessed care. Despite a substantial reduction in admissions during the strike period, we found no evidence that overall inpatient neonatal mortality changed. Although crude mortality increased from 12.5% to 14.9%, this difference was attenuated after accounting for hospital-level clustering. Interrupted time-series analyses that adjusted for hospital-specific trends and seasonality similarly found no evidence of a change in mortality rates. However, these findings should be interpreted with caution because the study included only 11 hospitals and was therefore unlikely to detect smaller mortality effects. In contrast, case fatality among neonates admitted with infection increased during the strike period, suggesting that strike-related disruptions may have disproportionately affected particularly vulnerable patient groups.

### Risk Selection and Case Mix Changes

The findings are consistent with a change in case mix during the strike period, with a greater proportion of admitted neonates potentially having more severe illness. One possible explanation is the prioritization of emergency admissions and restriction of non-urgent care during industrial action. A 2024 systematic review of healthcare worker strikes in sub-Saharan Africa identified service restriction and prioritization of emergency care as common mitigation strategies [32]. Similar shifts in case mix have been reported during the 2016 junior doctors’ strike in England [33]. In Kenya, a faith-based referral hospital that remained open during the 2016–2017 physicians’ strike reported restricting referrals and admissions in response to capacity constraints [13], illustrating mechanisms through which the composition of admitted patients may change during strike periods.

Routine data often miss deaths among the smallest newborns, making their recorded mortality look implausibly low. Using the NID, Malla and colleagues treated mortality of at least 700 per 1,000 among neonates under 1,000 g as a sign of adequate recording [17]. In our data, mortality among neonates under 1,000 g was 797 per 1,000 admissions before the strike and 944 during the strike, suggesting reasonably complete death recording among admitted neonates. Nevertheless, hospital mortality reflects outcomes only among patients who accessed care. During the strike, admissions decreased substantially, raising the possibility that the population reaching neonatal units differed from that seen before the strike. If the most vulnerable newborns were less likely to reach care, hospital-based mortality estimates may underestimate the overall mortality burden associated with neonatal illness during this period.

Inpatient mortality did not change measurably, but these estimates are restricted to newborns admitted to the study hospitals. An estimated 1,679 admissions (95% CI 329–3,029) did not occur during the strike. Any deaths among these newborns would have occurred outside the hospitals and were not measurable in this dataset. A qualitative study in Nigeria found health worker strikes were among the reasons women gave for giving birth at home, even where facility care was free [34]. Previous research suggests such missed admissions may be consequential: qualitative interviews on the effects of healthcare worker strikes in Kenya described newborn deaths and long-term complications from delayed treatment, and a quasi-experimental Kenyan study using strike exposure at birth estimated an additional 19 neonatal deaths per 1,000 births associated with the strike [35, 36]. Therefore, the true impact of the 2024 strike on neonatal mortality is likely underestimated by exclusive use of hospital-level data.

### Increase in Mortality Risk Attributed to Infection

During the strike, case fatality among neonates with infection as the primary discharge diagnosis doubled, from 4.7% to 9.6% (RR 2.06, 95% CI 1.18–3.58), while case fatality for prematurity and intrapartum-related conditions was unchanged. This increase may have been driven by families delaying presentation (or being refused admission) until a newborn’s condition was critical, or by an increase in hospital-acquired infections (HAIs). Refused admissions have been documented during previous Kenyan strikes: a faith-based referral hospital refused outside referrals and limited admission of high-risk pregnancies [13], and a public hospital turned away mothers when only life-threatening cases could be operated on [35]. Similar trends have been documented globally. For example, during a prolonged medics strike in Zimbabwe, widespread hospital closures similarly forced mothers to delay seeking care, directly exacerbating the severity of complications, including infection, and worsening clinical outcomes [37]. Many additional studies on neonatal care delays suggest that in the absence of accessible physicians, families often pursue self-medication, traditional healers, or herbalists, resulting in delayed diagnosis and treatment and poorer outcomes [38–40].

Additionally, we observed a case-mix shift toward outborn admissions within the severely reduced admissions hospitals group. Prior studies indicate that outborn neonates face a higher baseline infection risk due to unsafe delivery environments, delayed presentation, and prior exposure to suboptimal care [41]. Neonates admitted during the strike to hospitals with the steepest declines in admission also experienced prolonged hospitalization, with median length of stay rising from 4 days (IQR 2–8) to 9.5 days (IQR 4–25). While this is consistent with a higher-acuity case mix, it may also reflect slower clinical decision-making, including discharge decisions, in the absence of physicians. Prolonged hospitalization of this vulnerable population would have increased cumulative exposure to HAI risks, which is already substantial in resource-limited neonatal units and may therefore have contributed to the elevated infection case fatality observed in these hospitals [42].

Antibiotic-related processes may have further amplified this risk. Although not directly assessed in this study, physician absence may have disrupted appropriate antibiotic prescribing, ordering, and timely administration. Interruptions at any point in the medication-use process, including incorrect drug selection, delayed initiation, dosing errors, or missed opportunities for escalation, are well-documented drivers of deterioration in neonatal infections [43]. An Ethiopian neonatal intensive care unit study found that key risk factors for HAI for neonates included place of birth (outborn vs inborn), longer length-of-stay, and inappropriate antibiotic use; the first two increased during this strike period [44].

### Workload and Documented Care

None of the care measures evaluated changed significantly during the strike. Kenyan neonatal units report standard nurse-to-baby ratios of 1:13 to 1:25 [8–10], so with admissions significantly lower during the strike, nurses likely had more time per neonate. In the hospitals where admissions fell most steeply, coverage of several measures was higher during the strike. Pulse oximetry rose from 78.8% to 97.3%, phototherapy from 81.6% to 96.0%, and kangaroo mother care from 61.6% to 79.0%, while admission normothermia fell from 61.0% to 52.2% (Additional file 3). Nurse-led care may have been sustained through the disruption, consistent with the unchanged mortality per admission and in contrast with strikes in which nurses withdraw, where an analysis of New York State strikes between 1984 and 2004 found an 18% increase in in-hospital mortality [45].

### Strengths and Limitations

Strengths of this analysis include standardized data collection used consistently across all neonatal wards. The NEST360 NID dataset contributed a large volume of newborn records, enabling robust analysis of admissions. Additionally, the strike had an exactly dated onset and end, allowing both the disruption and the recovery to be estimated using ITS analysis, and admissions showed no detectable difference from seasonal baseline after the strike ended, which supports attribution of the utilization decline to the strike itself. The extended 72-week baseline allowed adjustment for secular trend and annual seasonality. The primary admissions estimate was significant across six model specifications, and comparisons across coverage measures, diagnoses, and birthweight bands were corrected for multiple testing within pre-specified families. This study is reported following the STROBE guideline (Additional file 6).

This study relied solely on inpatient data, so newborns who were never admitted (whether turned away, diverted to private or faith-based facilities, dying in transit, or never brought to care) are unobserved. Additionally, with only 11 hospital clusters, cluster-robust inference has limited precision despite the bias-reducing corrections applied, and analyses can detect only large effects. The single hospital that contracted temporary staff cannot be compared with the remaining ten in a way that separates its staffing response from its other characteristics. Hospital impact groups were defined by the strike-period admission decline itself, so group-level admission changes are, by construction, descriptive. Finally, it is important to note that these methods cannot fully separate increases in mortality attributable to changes in patient composition from those attributable to changes in the quality of care delivered.

Several features of these data indicate that documentation practice, rather than care alone, may have changed during the strike. The coverage analysis may also have lacked enough power to detect a true change as documented coverage varied substantially between hospitals for every measure. With eleven clusters, only large effects would be detectable. Within the strike period, completeness moved in opposite directions across variable types: recorded coverage of nurse-delivered interventions often held steady or increased, while variables captured at admission were recorded less completely. Birth location was unknown for 8.6% of pre-strike and 9.9% of strike-period admissions, and birthweight was unrecorded for 0.7% of pre-strike and 8.1% of strike-period admissions in the severely reduced admissions hospitals. Similarly, recorded neonatal infection as the primary discharge diagnosis is a clinical label without a standard definition, and previous work has shown gaps between documented and observed neonatal infection care [46].

### Mitigation and Health System Resilience

Protecting these populations from the negative effects of future healthcare disruptions requires building greater health system resilience against healthcare worker strikes [47]. An example of such resilience emerged within our dataset at one national referral hospital that contracted staff to maintain essential services. Previous research identifies comparable mitigation strategies put in place during other health system shocks. During the COVID-19 pandemic, hospitals addressed workforce shortages in newborn service delivery through task-sharing, locum and student staffing, virtual training, administrative support for staff, and telemedicine follow-up [48]. Similarly, during a 69-day residents’ strike in India, medical officers were retrained in labor-room and high-risk neonatal care [49]. The affected unit reported no significant differences in high-risk deliveries, admissions, or mortality across the observed periods [49].

### What Next in Research

Future research should aim to capture the broader impact of health worker strikes by including data on outcomes outside health facilities. Subsequent analyses could evaluate the effectiveness of mitigation strategies during strikes, examining how private and faith-based health networks absorb overflow from public systems, and determining which task-shifting models offer protective benefits without compromising patient safety. Preventing strikes is as important as mitigating them. Strikes in Kenya have generally arisen from long-standing workforce pressures, including staffing levels, remuneration, and working conditions [5, 35]. Sustained dialogue between government and health worker unions, equitable terms across cadres, and agreed mechanisms for resolving disputes may help reduce their occurrence. Because newborn care is predominantly nurse-delivered [8, 10], how strikes by different cadres affect neonatal and maternity services also warrants further study.

## Conclusions

The 2024 physicians’ strike in Kenya was associated with severe disruptions to neonatal service utilization. Among newborns who continued to reach care, hospital-level mortality per admission did not change measurably, but highly vulnerable subgroups were disproportionately affected. Case fatality among neonates with recorded infection doubled. Because this analysis was confined to inpatient events, outcomes of newborns who did not access hospitals were not captured. Both inpatient and population-level data are needed to fully assess the strike’s impact.

Drawing on the documented operational strategies noted previously, policymakers could establish broader frameworks to improve health system resilience against future service interruptions. Such frameworks might include formalizing preemptive contingency plans and establishing minimum service-level agreements. Contingency planning could incorporate the defined task-shifting protocols and telemedicine practices noted above, alongside the integration of faith-based and private providers into referral networks to establish baseline surge capacity [5, 13]. Implementing these structural measures may help maintain equitable access to care during periods of recurrent workforce disruption.

## Data Availability

Data sharing and transfer agreements were jointly developed and signed by all collaborating partners. The data may be available upon request from LSHTM data compass systems pending approval by a multi-country NEST360 Data Sharing and Publications Group. Data Tools are open access on www.NEST360.org and www.newborntoolkit.org. For the purpose of open access, the author has applied a 'Creative Commons Attribution' (CC BY) license to any Author Accepted Manuscript version arising.

## List of abbreviations

AR(1): First-order autoregressive
BH: Benjamini–Hochberg
CI: Confidence interval
CIN: Clinical Information Network
CPAP: Continuous positive airway pressure
GEE: Generalized estimating equation
HAI: Hospital acquired infection
IQR: Interquartile Range
IRR: Incidence rate ratio
ITS: Interrupted time series
KMC: Kangaroo Mother Care
LSHTM: London School of Hygiene & Tropical Medicine
NEST360: Newborn Essential Solutions and Technologies
NID: Neonatal Inpatient Dataset
OR: Odds ratio
QIC: Quasi-likelihood information criterion
RR: Risk ratio
SD: Standard deviation
SDG: Sustainable Development Goal
STROBE: Strengthening the Reporting of Observational Studies in Epidemiology
WHO: World Health Organization

## Declarations

### Ethics approval and consent to participate

NEST360 complex evaluation was granted ethical approval from the Institutional Review Boards of all four countries, the London School of Hygiene & Tropical Medicine (LSHTM) and Rice University. See Additional file 7 for a summary of relevant approvals.

### Consent for publication

Not applicable.

### Availability of data and materials

Data sharing and transfer agreements were jointly developed and signed by all collaborating partners. The data may be available upon request from LSHTM data compass systems pending approval by a multi-country NEST360 Data Sharing and Publications Group. Data Tools are open access on www.NEST360.org and www.newborntoolkit.org [50]. For the purpose of open access, the author has applied a ‘Creative Commons Attribution’ (CC BY) licence to any Author Accepted Manuscript version arising.

### Authors’ contributions

This analysis was done as part of ongoing work with the NEST360 Alliance and partners. MP, JW, and CAB developed the detailed research questions and overall analysis plan for this paper. Analysis was led by MP and supported by JW. MP, CM, JW, and CAB drafted the manuscript. WMM provided valuable guidance and support on the development of the manuscript. MP, CM, JW, GO, DRM, HM, DMM, FOO, JEL, NRR, RRK, ZMO, LRH, CAB, and WMM reviewed and helped to revise the manuscript. All authors reviewed and agreed on the final version. The authors’ views are their own, and not necessarily from any of the institutions they represent.

## Acknowledgements

We thank all the newborns and their families whose data were included in this analysis. We acknowledge the dedication of the healthcare workers across all study hospitals, whose commitment to newborn care, even amid challenging working conditions, made this research possible. We also recognize all hospital teams implementing with NEST360 and the data collection teams whose diligence underpins this work. James H Cross; Christine A Bohne; Lucas Malla; Morris Ondieki Ogero; John Wainaina; Eric O Ohuma; Samuel K Ngwala; Joseph Misyenje; Msandeni Chiume; Josephine Shabani; Irabi Kassim; Prosper Mshana; Jacqueline Minja; Honorati Masanja; Franklin Ochieng Okech; Olabisi Dosunmu; Rebecca E Penzias; Kristina Shemwell; Boniface Makone; Z Maria Oden; Rebecca Richards-Kortum; Henry Gathuri; Julius Thomas; Junwei Chen; Joy E Lawn.

## Additional files

**Additional file 1 (.docx).** Full ITS Model Output for Admissions and Mortality. Incidence rate ratios (95% CI) and p-values for each term of the primary segmented Poisson GEE models of weekly admissions and mortality.

**Additional file 2 (.docx).** Group-level Case-mix Detail by Admission Decline Group. Birthweight, prematurity, outborn share, and length of stay in the pre-strike and strike periods, by admission decline group.

**Additional file 3 (.docx).** Care Coverage by Admission Decline Group. Coverage of seven care processes in the pre-strike and strike periods, with 95% CIs and n/N, by admission decline group.

**Additional file 4 (.docx).** Case-Fatality Rate by Admission Decline Group. Case fatality for infection, prematurity, and intrapartum-related conditions in the pre-strike and strike periods, with 95% CIs, by admission decline group.

**Additional file 5 (.docx).** Birthweight Band Mortality by Admission Decline Group. Inpatient mortality by birthweight band in the pre-strike and strike periods, with 95% CIs, by admission decline group.

**Additional file 6 (.docx).** Strengthening the Reporting of Observational Studies in Epidemiology (STROBE) Checklist. Completed STROBE checklist with manuscript page and line numbers for each item.

**Additional file 7 (.docx).** Local ethical approval for the complex evaluation of the implementation of a small and sick newborn care package with NEST360. Local ethics committee approvals in each country, with protocol titles and approval reference numbers.

