## Additional file 1 for "Physicians’ Industrial Action: An Interrupted Time Series Analysis Assessing Impact in 11 Kenyan Neonatal Units"

### Additional file 1: Full ITS Model Output for Admissions and Mortality

| **Model term** | **IRR (95% CI)** | ***p*** |
|---|---|---|
| *Admissions (exchangeable working correlation, primary specification)* | | |
| Baseline trend (time) | 1.000 (0.997–1.003) | 0.847 |
| Strike-period level change (during) | 0.485 (0.313–0.753) | 0.004 |
| Post-strike level (post) | 1.000 (0.814–1.229) | 0.999 |
| Post-strike slope (post_time) | 1.003 (0.993–1.013) | 0.502 |
| *Mortality (exchangeable working correlation, primary specification)* | | |
| Baseline trend (time) | 1.001 (0.992–1.011) | 0.736 |
| Strike-period level change (during) | 0.877 (0.531–1.449) | 0.574 |
| Post-strike level (post) | 0.852 (0.493–1.473) | 0.530 |
| Post-strike slope (post_time) | 0.986 (0.948–1.027) | 0.461 |
