## Additional file 2 for "Physicians’ Industrial Action: An Interrupted Time Series Analysis Assessing Impact in 11 Kenyan Neonatal Units"

### Additional file 2: Group-level Case-mix Detail by Admission Decline Group

| Characteristic | Pre-strike | Strike |
|---|---|---|
| *Severely reduced* | | |
| Birthweight, mean ± SD (*n*) | 2,677 ± 870 (2,278) | 2,138 ± 901 (171) |
| Prematurity, *n/N* (%) | 726/2,294 (31.6%) | 101/186 (54.3%) |
| Outborn, *n/N* (%) | 433/2,294 (18.9%) | 106/186 (57.0%) |
| Length of stay, median [Q1–Q3] (*n*) | 4 [2–8] (2,290) | 10 [4–25] (186) |
| *Moderately reduced* | | |
| Birthweight, mean ± SD (*n*) | 2,682 ± 848 (2,452) | 2,665 ± 853 (997) |
| Prematurity, *n/N* (%) | 825/2,469 (33.4%) | 362/1,010 (35.8%) |
| Outborn, *n/N* (%) | 277/2,469 (11.2%) | 142/1,010 (14.1%) |
| Length of stay, median [Q1–Q3] (*n*) | 5 [3–10] (2,455) | 5 [2–9] (1,010) |
| *Minimally reduced* | | |
| Birthweight, mean ± SD (*n*) | 2,564 ± 906 (595) | 2,401 ± 872 (309) |
| Prematurity, *n/N* (%) | 273/601 (45.4%) | 187/316 (59.2%) |
| Outborn, *n/N* (%) | 97/601 (16.1%) | 57/316 (18.0%) |
| Length of stay, median [Q1–Q3] (*n*) | 4 [2–8] (600) | 5 [2–10] (316) |
