## Additional file 3 for "Physicians’ Industrial Action: An Interrupted Time Series Analysis Assessing Impact in 11 Kenyan Neonatal Units"

### Additional file 3: Care Coverage by Admission Decline Group

| **Measure** | **Pre-strike % (95% CI)** | **Pre-strike *n/N*** | **Strike % (95% CI)** | **Strike *n/N*** |
|---|---|---|---|---|
| *Severely reduced* | | | | |
| Glucose screening | 84.9 (83.4–86.3) | 1,948/2,294 | 93.5 (89.1–96.3) | 174/186 |
| Pulse oximetry | 78.8 (77.0–80.4) | 1,807/2,294 | 97.3 (93.9–98.8) | 181/186 |
| Temp. screening | 93.2 (92.1–94.2) | 2,139/2,294 | 96.8 (93.1–98.5) | 180/186 |
| KMC (1000-1999g) | 61.6 (57.3–65.8) | 302/490 | 79.0 (68.9–86.5) | 64/81 |
| Phototherapy (jaundice) | 81.6 (77.2–85.4) | 280/343 | 96.0 (86.5–98.9) | 48/50 |
| CPAP (eligible) | 25.2 (20.2–30.9) | 63/250 | 23.8 (13.5–38.5) | 10/42 |
| Normothermia | 61.0 (58.9–63.1) | 1,305/2,139 | 52.2 (45.0–59.4) | 94/180 |
| *Moderately reduced* | | | | |
| Glucose screening | 76.3 (74.6–78.0) | 1,885/2,469 | 74.6 (71.8–77.1) | 753/1,010 |
| Pulse oximetry | 80.8 (79.2–82.3) | 1,994/2,469 | 82.9 (80.4–85.1) | 837/1,010 |
| Temp. screening | 96.4 (95.6–97.1) | 2,381/2,469 | 95.2 (93.8–96.4) | 962/1,010 |
| KMC (1000-1999g) | 60.5 (56.2–64.7) | 308/509 | 62.4 (55.4–68.9) | 121/194 |
| Phototherapy (jaundice) | 82.4 (78.9–85.4) | 426/517 | 82.1 (75.3–87.3) | 128/156 |
| CPAP (eligible) | 51.7 (45.4–57.9) | 124/240 | 44.4 (34.6–54.7) | 40/90 |
| Normothermia | 53.0 (51.0–55.0) | 1,261/2,381 | 52.3 (49.1–55.4) | 503/962 |
| *Minimally reduced* | | | | |
| Glucose screening | 92.2 (89.8–94.1) | 554/601 | 85.1 (80.8–88.6) | 269/316 |
| Pulse oximetry | 92.8 (90.5–94.6) | 558/601 | 94.0 (90.8–96.1) | 297/316 |
| Temp. screening | 91.3 (88.8–93.3) | 549/601 | 93.4 (90.1–95.6) | 295/316 |
| KMC (1000-1999g) | 69.6 (61.4–76.6) | 96/138 | 74.0 (64.6–81.6) | 74/100 |
| Phototherapy (jaundice) | 66.7 (53.0–78.0) | 34/51 | 83.3 (64.1–93.3) | 20/24 |
| CPAP (eligible) | 58.7 (47.4–69.1) | 44/75 | 48.7 (33.9–63.8) | 19/39 |
| Normothermia | 46.1 (42.0–50.3) | 253/549 | 47.1 (41.5–52.8) | 139/295 |
