## Additional file 4 for "Physicians’ Industrial Action: An Interrupted Time Series Analysis Assessing Impact in 11 Kenyan Neonatal Units"

### Additional file 4: Case-Fatality Rate by Admission Decline Group

| **Diagnosis** | **Pre-strike *n/N* (%) [95% CI]** | **Strike *n/N* (%) [95% CI]** |
|---|---|---|
| *Severely reduced* | | |
| Infection | 16/316 (5.1%) [3.1–8.1] | 6/27 (22.2%) [10.6–40.8] |
| Prematurity | 101/726 (13.9%) [11.6–16.6] | 17/101 (16.8%) [10.8–25.3] |
| Intrapartum-related | 91/603 (15.1%) [12.5–18.2] | 4/26 (15.4%) [6.2–33.5] |
| *Moderately reduced* | | |
| Infection | 18/510 (3.5%) [2.2–5.5] | 12/161 (7.5%) [4.3–12.6] |
| Prematurity | 153/825 (18.5%) [16.0–21.3] | 73/362 (20.2%) [16.4–24.6] |
| Intrapartum-related | 107/562 (19.0%) [16.0–22.5] | 49/273 (17.9%) [13.9–22.9] |
| *Minimally reduced* | | |
| Infection | 11/134 (8.2%) [4.6–14.1] | 3/30 (10.0%) [3.5–25.6] |
| Prematurity | 70/273 (25.6%) [20.8–31.1] | 34/187 (18.2%) [13.3–24.3] |
| Intrapartum-related | 30/115 (26.1%) [18.9–34.8] | 15/48 (31.2%) [19.9–45.3] |
