## Additional file 5 for "Physicians’ Industrial Action: An Interrupted Time Series Analysis Assessing Impact in 11 Kenyan Neonatal Units"

### Additional file 5: Birthweight Band Mortality by Admission Decline Group

| **Birthweight band** | **Pre-strike *n/N* (%) [95% CI]** | **Strike *n/N* (%) [95% CI]** |
|---|---|---|
| *Severely reduced* | | |
| 400–999 g | 43/60 (71.7%) [59.2–81.5] | 8/8 (100.0%) [67.6–100.0] |
| 1,000–1,499 g | 62/178 (34.8%) [28.2–42.1] | 8/40 (20.0%) [10.5–34.8] |
| 1,500–1,999 g | 28/312 (9.0%) [6.3–12.7] | 6/41 (14.6%) [6.9–28.4] |
| 2,000–2,499 g | 21/321 (6.5%) [4.3–9.8] | 1/18 (5.6%) [1.0–25.8] |
| ≥2,500 g | 98/1,404 (7.0%) [5.8–8.4] | 4/64 (6.2%) [2.5–15.0] |
| *Moderately reduced* | | |
| 400–999 g | 51/59 (86.4%) [75.5–93.0] | 33/36 (91.7%) [78.2–97.1] |
| 1,000–1,499 g | 58/171 (33.9%) [27.2–41.3] | 27/76 (35.5%) [25.7–46.7] |
| 1,500–1,999 g | 43/338 (12.7%) [9.6–16.7] | 12/118 (10.2%) [5.9–16.9] |
| 2,000–2,499 g | 28/331 (8.5%) [5.9–12.0] | 9/125 (7.2%) [3.8–13.1] |
| ≥2,500 g | 108/1,552 (7.0%) [5.8–8.3] | 60/642 (9.3%) [7.3–11.8] |
| *Minimally reduced* | | |
| 400–999 g | 16/19 (84.2%) [62.4–94.5] | 10/10 (100.0%) [72.2–100.0] |
| 1,000–1,499 g | 21/46 (45.7%) [32.2–59.8] | 12/30 (40.0%) [24.6–57.7] |
| 1,500–1,999 g | 24/92 (26.1%) [18.2–35.9] | 10/70 (14.3%) [7.9–24.3] |
| 2,000–2,499 g | 13/106 (12.3%) [7.3–19.9] | 1/49 (2.0%) [0.4–10.7] |
| ≥2,500 g | 40/332 (12.0%) [9.0–16.0] | 19/150 (12.7%) [8.3–18.9] |
