## Additional file 6 for "Physicians’ Industrial Action: An Interrupted Time Series Analysis Assessing Impact in 11 Kenyan Neonatal Units"

### Additional file 6: Strengthening the Reporting of Observational Studies in Epidemiology (STROBE) Checklist

Page and line numbers refer to the main manuscript.

| Section/Topic | Item No. | Recommendation | Page No. (lines) | Relevant text from manuscript |
|---|---|---|---|---|
| **Title and abstract** | | | | |
| **Title and abstract** | 1 | (a) Indicate the study's design with a commonly used term in the title or the abstract | p. 1, L1–2<br>p. 2, L37 | Title: "Physicians' Industrial Action: An Interrupted Time Series Analysis…"<br>Abstract: "We conducted a retrospective interrupted time-series analysis…" |
| | | (b) Provide in the abstract an informative and balanced summary of what was done and what was found | pp. 2–3, L32–59 | Structured abstract (Background, Methods, Results, Conclusion) reporting the admissions decline (IRR 0.49, 95% CI 0.31–0.75), no detectable change in care coverage or overall mortality, and the doubling of infection case fatality. |
| **Introduction** | | | | |
| **Background/rationale** | 2 | Explain the scientific background and rationale for the investigation being reported | pp. 4–5, L62–95 | Recurrent Kenyan health-worker strikes and neonatal burden (L62–68); staffing constraints and physician role in neonatal units (L69–77); gap in neonatal-specific evidence (L78–82); the 2024 strike and NEST360 data as an opportunity (L83–95). |
| **Objectives** | 3 | State specific objectives, including any prespecified hypotheses | p. 6, L96–102<br>p. 5, L75–77 | Changes in (1) admissions, (2) care practices, and (3) inpatient outcomes (L96–102).<br>Expected direction of effect: strike "would therefore be expected to disproportionately compromise care for the sickest and most complex neonatal admissions" (L75–77). |
| **Methods** | | | | |
| **Study design** | 4 | Present key elements of study design early in the paper | p. 6, L106–110 | Study Design and Setting: retrospective, multicenter interrupted time series analysis of 11 neonatal units implementing with NEST360. |
| **Setting** | 5 | Describe the setting, locations, and relevant dates, including periods of recruitment, exposure, follow-up, and data collection | pp. 6–7, L106–120<br>p. 7, L122–125 | 11 public hospital neonatal units in Kenya (1 Level III, 10 Level II with CPAP) (L106–110). Strike 15 Mar–9 May 2024; pre-strike 15 Dec 2023–14 Mar 2024; post-strike 10 May–9 Aug 2024; extended ITS window 20 Mar 2023–4 Aug 2024 (L111–120). Data abstracted from routine inpatient records by trained data officers (L122–125). |

| Section/Topic | Item No. | Recommendation | Page No. (lines) | Relevant text from manuscript |
|---|---|---|---|---|
| **Participants** | 6 | (a) Cohort study—Give the eligibility criteria, and the sources and methods of selection of participants. Describe methods of follow-up | p. 6, L106–109<br>p. 7, L126–131<br>p. 7, L132–134 | Admissions to 11 of 13 Kenyan NEST360 public hospitals; two hospitals excluded for unrelated pre-strike disruptions (L106–109). Admissions within the three periods form the descriptive cohort (L126–131). Follow-up to discharge outcome; unknown/transfer outcomes excluded from mortality denominators (L132–134). |
| | | (b) Cohort study—For matched studies, give matching criteria and number of exposed and unexposed | N/A | Not a matched study. |
| **Variables** | 7 | Clearly define all outcomes, exposures, predictors, potential confounders, and effect modifiers. Give diagnostic criteria, if applicable | p. 6, L111–115<br>pp. 7–8, L137–155<br>p. 9, L166, L173–174 | Exposure: strike period (L111–115). Primary outcomes: weekly admissions and weekly inpatient mortality; secondary variables and eligibility criteria for each coverage measure; normothermia 36.5–37.5 °C (L142–149). Infection diagnosis definition (L137–140). Impact groups by admission decline (L150–155). Secular trend and seasonality (harmonic terms) as adjustment variables (L166, L173–174). |
| **Data sources/ measurement** | 8* | For each variable of interest, give sources of data and details of methods of assessment (measurement). Describe comparability of assessment methods if there is more than one group | p. 7, L122–125<br>p. 7, L132–135<br>p. 18, L377–378 | NEST360 Neonatal Inpatient Dataset, standardized and abstracted through Kenya's CIN (L122–125). Mortality from discharge status; admission type from birth location; diagnosis from primary discharge diagnosis (L132–135). Standardized data collection used consistently across all units (L377–378). |
| **Bias** | 9 | Describe any efforts to address potential sources of bias | p. 6, L108–109<br>pp. 6–7, L116–120<br>p. 8, L154–155<br>p. 10, L188–190<br>p. 15, L310–314 | Exclusion of hospitals with unrelated disruptions (L108–109); extended window for seasonality and per-hospital screening for other disruptions (L116–120); outcome-defined groups reported descriptively only (L154–155); bias-reduced small-cluster standard errors (L188–190); check for under-ascertainment of deaths <1,000 g (L310–314). See also limitations (pp. 18–19, L387–408). |
| **Study size** | 10 | Explain how the study size was arrived at | p. 7, L126–131<br>p. 11, L208–213 | All admissions in the 11 units over the analysis window were used: 27,638 for ITS models; 12,184 in the three study periods. |
| **Quantitative variables** | 11 | Explain how quantitative variables were handled in the analyses. If applicable, describe which groupings were chosen and why | p. 8, L143–153<br>p. 8, L157–158; p. 9, L173–174<br>p. 22, Table 3 | Birthweight thresholds defining KMC/CPAP eligibility; normothermia range; impact groups split at the median decline (L143–153). Admissions aggregated to unit-weeks; time modeled with trend and Fourier terms (L157–158, L173–174). Birthweight bands (400–999 g to ≥2,500 g) shown in Table 3. |

| Section/Topic | Item No. | Recommendation | Page No. (lines) | Relevant text from manuscript |
|---|---|---|---|---|
| **Statistical methods** | 12 | (a) Describe all statistical methods, including those used to control for confounding | pp. 8–10, L157–204 | Segmented Poisson regression with GEE clustered by unit; secular trend and harmonic seasonal terms; log offset for mortality; logistic/log GEE for coverage and case fatality; QIC-selected working correlation for admissions and mortality, exchangeable for coverage and case fatality; Mancl–DeRouen correction with t(10) inference; Benjamini–Hochberg within pre-specified families; Python 3.12. |
| | | (b) Describe any methods used to examine subgroups and interactions | p. 8, L150–155<br>p. 9, L179–183; p. 10, L196–197 | Stratification by admission decline (descriptive only) (L150–155); diagnosis- and birthweight-specific comparisons (L179–183); hospital-level segmented models (L196–197). |
| | | (c) Explain how missing data were addressed | p. 7, L132–134<br>p. 9, L177–178 | Unknown/transfer outcomes excluded from mortality denominators (L132–134); unit-weeks with zero analyzable admissions excluded (n = 6) (L177–178). |
| | | (d) Cohort study—If applicable, explain how loss to follow-up was addressed | p. 7, L132–134 | Admissions with transfer or unknown outcome retained in admission counts but excluded from mortality denominators. |
| | | (e) Describe any sensitivity analyses | p. 10, L192–198 | Six sensitivity analyses: negative binomial GEE; 3-month baseline without harmonics; strike-anchored weeks; pooled negative binomial with Newey–West SEs; hospital-level models; AR(1) working correlation. |
| **Results** | | | | |
| **Participants** | 13* | (a) Report numbers of individuals at each stage of study—eg numbers potentially eligible, examined for eligibility, confirmed eligible, included in the study, completing follow-up, and analysed | p. 7, L126–131<br>p. 7, L134<br>p. 11, L208–213 | 27,638 admissions in the ITS window; 12,184 in study periods (5,364 pre-strike, 1,512 strike, 5,308 post-strike); 12,175 with known outcome; 1,501 deaths. |
| | | (b) Give reasons for non-participation at each stage | p. 6, L106–109<br>p. 7, L132–134 | Two of 13 hospitals excluded for unrelated pre-strike disruptions; admissions with unknown/transfer outcomes excluded from mortality analyses. |
| | | (c) Consider use of a flow diagram | — | N/A |
| **Descriptive data** | 14* | (a) Give characteristics of study participants (eg demographic, clinical, social) and information on exposures and potential confounders | p. 11, L214–227; p. 21, Table 1<br>Additional file 2 | Table 1: admission type, birthweight, primary diagnosis and mortality by period; text summary of case-mix shift (L214–227); group-level detail in Additional file 2. |

| Section/Topic | Item No. | Recommendation | Page No. (lines) | Relevant text from manuscript |
|---|---|---|---|---|
| | | (b) Indicate number of participants with missing data for each variable of interest | p. 21, Table 1; p. 11, L219–221<br>p. 19, L404–405 | Table 1 reports unknown admission type, birthweight n, and "not recorded" diagnosis; birth location unknown by period (L219–221); unrecorded birthweight in the severely reduced group (L404–405). |
| | | (c) Cohort study—Summarise follow-up time (eg, average and total amount) | p. 11, L211–213<br>p. 11, L225–226 | 374 unit-weeks (descriptive window) and 792 unit-weeks (ITS window); median length of stay reported for the severely reduced group only. |
| **Outcome data** | 15* | Cohort study—Report numbers of outcome events or summary measures over time | p. 11, L210–211<br>p. 12, L229–231<br>p. 13, L268–270<br>p. 14, L273–280<br>p. 22, Table 3 | 1,501 deaths among 12,175 with known outcome; weekly admissions by period; infection deaths 45/960 vs 21/218; deaths by period and birthweight band (Table 3). |
| **Main results** | 16 | (a) Give unadjusted estimates and, if applicable, confounder-adjusted estimates and their precision (eg, 95% confidence interval). Make clear which confounders were adjusted for and why they were included | p. 12, L231–236<br>p. 12, L244–251<br>p. 22, Table 2<br>p. 13, L268–270<br>p. 14, L276–280<br>p. 22, Table 3 | Admissions IRR 0.49 (95% CI 0.31–0.75); care coverage with unadjusted CIs (Table 2); crude mortality 12.5% vs 14.9% (RR 1.07, 0.85–1.33) and trend- and seasonality-adjusted IRR 0.88 (0.53–1.45); infection CFR RR 2.06 (1.18–3.58). Adjustment for secular trend and seasonality explained in Methods (p. 6, L116–118; p. 9, L166, L173–174). |
| | | (b) Report category boundaries when continuous variables were categorized | p. 8, L143–153<br>p. 22, Table 3 | Birthweight eligibility thresholds; group boundaries (55.7–96.5% and 20.8–50.5% decline); birthweight bands in Table 3. |
| | | (c) If relevant, consider translating estimates of relative risk into absolute risk for a meaningful time period | p. 12, L244–247<br>p. 13, L268–270<br>p. 14, L274–275 | Absolute shortfall of 1,679 admissions (95% CI 329–3,029); CFR 4.7% vs 9.6%; mortality per 1,000 by birthweight band. |
| **Other analyses** | 17 | Report other analyses done—eg analyses of subgroups and interactions, and sensitivity analyses | p. 12, L232–236<br>p. 12, L237–243; p. 24, Figure 2<br>p. 11, L222–227<br>p. 13, L260–266<br>Additional files 1–5 | Sensitivity estimates (IRR 0.43–0.64); hospital-level IRRs; group-level case mix and coverage (descriptive); full model output and group-level tables in Additional files. |
| **Discussion** | | | | |
| **Key results** | 18 | Summarise key results with reference to study objectives | pp. 14–15, L285–298 | Summary of admissions decline, case-mix shift, no measurable mortality change, and increased infection case fatality. |

| Section/Topic | Item No. | Recommendation | Page No. (lines) | Relevant text from manuscript |
|---|---|---|---|---|
| **Limitations** | 19 | Discuss limitations of the study, taking into account sources of potential bias or imprecision. Discuss both direction and magnitude of any potential bias | pp. 18–19, L387–408<br>p. 14, L294–295<br>p. 15, L317–319<br>p. 16, L329–330 | Inpatient-only data; 11 clusters and limited precision; outcome-defined groups; case mix vs quality of care; documentation changes (L387–408). Direction of bias: selective non-admission of the most fragile newborns would be invisible in the data and would bias birthweight-specific mortality (L317–319); total impact likely underestimated (L329–330). |
| **Interpretation** | 20 | Give a cautious overall interpretation of results considering objectives, limitations, multiplicity of analyses, results from similar studies, and other relevant evidence | pp. 15–18, L300–375<br>p. 18, L384–386<br>pp. 19–20, L410–419<br>pp. 20–21, L433–446 | Interpretation of risk selection, infection CFR, and documented care with comparison to prior strike studies (L300–375); multiplicity correction noted (L384–386); mitigation and health system resilience (L410–419); cautious conclusions and implications (L433–446). |
| **Generalisability** | 21 | Discuss the generalisability (external validity) of the study results | p. 6, L106–110<br>pp. 18–19, L391–392 | Setting limited to 11 NEST360-supported public hospitals; single hospital with contracted staff not comparable. |
| **Other information** | | | | |
| **Funding** | 22 | Give the source of funding and the role of the funders for the present study and, if applicable, for the original study on which the present article is based | p. 27, L514–518 | Funded through the NEST360 Alliance (Gates Foundation, ELMA Philanthropies, Mohamed bin Zayed Foundation for Humanity, Beginnings Fund, Sall Family Foundation, private donors; agreements with Rice University). |

**Give information separately for cases and controls in case-control studies and, if applicable, for exposed and unexposed groups in cohort and cross-sectional studies.*
