## Additional file 7 for "Physicians’ Industrial Action: An Interrupted Time Series Analysis Assessing Impact in 11 Kenyan Neonatal Units"

**Additional file 7: Local ethical approval for the complex evaluation of the implementation of a small and sick newborn care package with NEST360**

| Country | Protocol Title | LEC Protocol ID |
|---|---|---|
| **Kenya** | Using a Health Facility Assessment to Assess Quality of Newborn Care in Kenya | MSU/DRPI/MUERC/00810/19 |
| **Malawi** | Using a Health Facility Assessment to Assess Quality of Newborn Care in Malawi | NHSRC 2463 |
| **Nigeria** | Quality Improvement Study of the Implementation of a Package of Trainings and Technologies for the Delivery of Comprehensive Newborn Care in Nigeria: A Multi-Country Study | **LUTH:** ADM/DCST/HREC/APP/3487<br>**UCH:** UI/EC/20/0713<br>**NHREC:** NHREC/01/01/2007 |
| **Tanzania** | Implementation study to improve the quality of comprehensive newborn care through introduction of the package of Newborn Essential Solutions and Technologies (NEST) in Tanzania | **IHI:** IHI/IRB/01-2021<br>**MUHAS:** MUHAS-REC-12-2019-072<br>**NIMR:** 3405 |

**Abbreviations**: LEC; Local Ethics Committee, ID; Identity, MSU; Michigan State University, DRPI; Disability Right Promotion International, MUERC; Maseno University Ethics Review

Committee, NHSRC; National Health Science Research Committee, LUTH; Lagos University Teaching Hospital, UCH; University College Hospital, NHREC; National Health Research Ethics Committee, IHI; Ifakara Health Institute, MUHAS; Muhimbili University of Health and Allied Science, NIMR; National Institute for Medical Research.
